# Relationship between circulating FSH levels and body composition and bone health in patients with prostate cancer who undergo androgen deprivation therapy. The BLADE study

**DOI:** 10.1101/2023.10.24.23297490

**Authors:** M Bergamini, A Dalla Volta, C Palumbo, S Zamboni, L Triggiani, M Zamparini, M Laganà, L Rinaudo, N Di Meo, I Caramella, R Bresciani, F Valcamonico, P Borghetti, A Guerini, D Farina, A Antonelli, C Simeone, G Mazziotti, A Berruti

## Abstract

**Background:** Amongst its extragonadal effects, Follicle-Stimulating Hormone (FSH) has an impact on body composition and bone metabolism. Since androgen deprivation therapy (ADT) has a profound impact on circulating FSH concentrations, this hormone could potentially be implicated in the changes of fat body mass (FBM), lean body mass (LBM) and bone fragility induced by ADT. The objective of this study is to correlate FSH serum levels with body composition parameters, bone mineral density (BMD) and bone turnover markers at baseline conditions and after 12 months of ADT.

**Methods:** 29 consecutive non-metastatic prostate cancer (PC) patients were enrolled from 2017 to 2019 in a phase IV study. All patients underwent administration of the Luteinizing Hormone-Releasing Hormone antagonist degarelix. FBM, LBM and BMD were evaluated by dual-energy x-ray absorptiometry at baseline and after 12 months of ADT. FSH, alkaline phosphatase (ALP) and C-terminal telopeptide of type I collagen (CTX) were assessed at baseline and after 6 and 12 months. For outcome measurements and statistical analysis, T-test or sign test and Pearson or Spearman tests for continuous variables were used when indicated.

**Results:** At baseline conditions, a weak, non-significant, direct relationship was found between FSH serum levels and FBM at arms (r=0.36) and legs (r=0.33). Conversely, a stronger correlation was observed between FSH and total FBM (r=0.52, p=0.006), fat mass at arms (r=0.54, p=0.004) and fat mass at trunk (r=0.45, p=0.018) assessed after 12 months. On the other hand, an inverse relationship between serum FSH and appendicular lean mass index (ALMI)/FBM ratio was observed (r=0.64, p=0.001). This is an ancillary study of a prospective trial and this is the main limitation.

**Conclusions:** FSH serum levels after ADT could have an impact on body composition, in particular on fat body mass. Therefore, FSH could be a promising marker to monitor the risk of sarcopenic obesity and cardiovascular complications in PC patients undergoing ADT.

**Funding:** this research was partially funded by Ferring Pharmaceuticals. The funder had no role in design and conduct of the study, collection, management, analysis, and interpretation of the data and in preparation, review, or approval of the manuscript.

**Clinical trial number:** clinicalTrials.gov NCT03202381, EudraCT Number 2016-004210-10.

## Introduction

Recent preclinical and clinical studies have shown that Follicle-Stimulating Hormone (FSH) exerts effects beyond those on gonadal tissue, which have been known for a long time (Lizneva et al. 2019). In the animal model, FSH has been observed to stimulate osteoclastic activity and therefore to exert a negative role on bone mass (Sun et al. 2006; Zaidi et al. 2023; Gera et al. 2022). Furthermore, FSH was found to have a positive action on adipocytes and the blockade of FSH induced by antibodies against FSH receptor caused an increase in bone mass and a reduction of body fat (Liu et al. 2017; Rojekar et al. 2023). These preclinical data were confirmed in clinical studies. Large epidemiologic data have shown significant reductions in bone mineral density (BMD) and high resorption rates ∼2-3 years prior to menopause, when FSH serum levels are increasing, which is also associated with increased body weight and visceral adiposity (Thurston et al.

2009; Senapati et al. 2014). A longitudinal study involving post-menopausal women has shown that increases in circulating FSH levels were associated with greater increases in the percentage of total body fat, total body fat mass and subcutaneous adipose tissue (Mattick et al. 2022). Moreover, clinical studies have found that FSH is also involved in the modulation of lean mass. A large cohort study of perimenopausal women showed that increased FSH concentrations were associated with increasing fat mass and decreasing lean mass measured by bioelectrical impedance analysis (Sowers et al. 2007). Another recent prospective study showed that the indicators of sarcopenia were strongly associated with gonadotropins levels, especially in older men (Guligowska et al. 2021). The impact of FSH on body composition may at least partially explain the observed correlation between FSH levels and the risk of cardiovascular events (Munir et al. 2012; Celestino Catão Da Silva et al. 2013; El Khoudary et al. 2016).

Studies clarifying the relationship among FSH, body composition measures and BMD in prostate cancer (PC) patients are lacking. Likewise, the interaction between the variation of FSH serum levels and the modification of these parameters in PC patients undergoing androgen deprivation therapy (ADT) is not known.

We recently conducted the BLADE study (Bone mineraL mAss Dexa dEgarelix), a prospective phase IV study designed to obtain explorative information on dual-energy x-ray absorptiometry (DXA) measurement changes in lean body mass (LBM) and fat body mass (FBM) in patients with non-metastatic PC treated with degarelix. Data on changes in total LBM and FBM and on the impact of degarelix on BMD and bone turnover markers were recently published (Palumbo et al. 2021; Palumbo et al. 2022).

Here we analyse the association of FSH with DXA-assessed FBM, LBM and BMD and bone turnover markers at baseline and after degarelix administration in the patients enrolled in the BLADE study.

## Patients and methods

### Trial design and endpoints

BLADE is a single-centre, prospective, interventional phase IV study (clinicalTrials.gov NCT03202381, EudraCT Number 2016-004210-10) conducted at the Prostate Cancer Unit of the Azienda Socio Sanitaria Territoriale degli Spedali Civili and Università degli Studi of Brescia. The study was carried out in accordance with the Declaration of Helsinki Principles and Good Clinical Practices and was approved by the Ethics Committee of Brescia (approval number NP2540). All patients provided a written informed consent. Male patients with histologically confirmed PC without bone metastasis at bone scintigraphy, judged eligible to ADT according to current guidelines recommendations (Cornford et al. 2017; Mottet et al. 2017) after a multidisciplinary discussion, were enrolled. Eligibility criteria have been published elsewhere (Palumbo, et al. 2021). Degarelix was administered as a subcutaneous injection with a starting dose of 240 mg, followed by a maintenance dose of 80 mg every 28 days. After 12 months, treatment with degarelix was continued as clinically indicated. DXA measurements for assessing BMD and body composition parameters were performed at baseline and after 12 months, using Hologic QDR-4500W instrumentation (Hologic Corporation, Waltham, Massachusetts).

### Assessment of regional lean body mass and fat body mass by dual x-ray absorptiometry

DXA measurements related to whole body DXA scans were extracted from Apex Software version 3.4.

The densitometric image of each patient was divided, following the manufacturer’s instructions, into different body districts, including arms, legs, trunk, head and other derived regions, such as the android and gynoid zone.

BMD, bone mineral content (BMC), Fat Free Mass and Fat mass were assessed for every region of interest, where Fat Free Mass was provided by the software in terms of lean soft tissue plus BMC. Despite the lean mass measured by DXA counts also skin, connective tissue and some lean components within the adipose tissue (Visser et al. 1999), it still correlates highly with computed tomography and magnetic resonance imaging measurements and represents a good approximation of the real muscle mass (Buckinx et al. 2018).

Other DXA derived body composition parameters, such as fat mass index (FM/H2) (FMI), appendicular lean mass index (ALM/H2) (ALMI) and Trunk/Appendicular ratio, were then calculated to complete the analysis and the patient characterization.

### Circulating bone turnover markers

Blood chemistry and bone turnover markers: alkaline phosphatase (ALP) and C-terminal telopeptide of type I collagen (CTX) were assessed at baseline, 6 and 12 months. CTX serum levels were measured using the ElectroChemiLuminescenceAssay (ECLIA) kit Elecsys beta-CrossLaps/serum (Roche Diagnostic, Germany) using Cobas e411instruments (Roche); normal ranges were < 0.704 ng/ml (men of the age between 50-70), < 0.854 ng/ml (men > 70) with a repeatability CV% of 2.6. Bone-ALP serum levels were determined in a twostep procedure. Briefly, total ALP serum activity was measured using the colorimetric method ALP2 (Roche) using Cobas c701 instruments (Roche); normal ranges were 50-116 U/L +/- 0.6 with a repeatability CV% of 0.7. Samples were then subjected to electrophoretic separation to separate the different ALP isoforms using the G26 automated system (Sebia, France) equipped with the Interlab specific kit (Italy).

Bone-ALP activity was calculated as fraction of the total ALP activity related to the percentage of densitometric analysis of the electrophoretic migration

### Statistical analysis

The normal distribution of continuous variables was tested with the Shapiro-Wilk test. Differences between parameters at baseline and 12 months were computed as percentage changes. To test if these changes were significantly different from 0 we used one sample t-test, or alternatively the non-parametric sign test.

Correlations between variables either at baseline and 12 months were expressed as Pearson’s r (or alternatively with Spearman R, for variables not normally distributed).

We considered a significant threshold of p < 0.05, and to control for possible false positive results we applied the Bonferroni correction (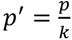, where k is the number of hypotheses tested). Given that Type I errors cannot decrease (the whole point of Bonferroni adjustments) without inflating type II errors (Perneger 1998), significant results in the raw test but which do not remain so after correction will still be mentioned.

Statistics were performed using R and SPSS (IBM Corp. Released 2015. IBM SPSS Statistics for Windows, Version 23.0. Armonk, NY: IBM Corp.)

## Results

Twenty-nine patients were included in the BLADE study and their characteristics have been described elsewhere (Palumbo, et al. 2022). The consort diagram is reported in supplementary material (Figure 1S). Mean FSH serum levels at baseline conditions were 11.9 UI/L (95% Confidence Intervals [CIs]: 7.6 to 16.3). Mean FSH serum levels significantly decreased to 1.45 UI/L (95% CIs: 1.01 to 1.89) after 6 months of degarelix treatment and to 2.4 UI/L (95% CIs: 1.4 to 3.4) after 12 months. The corresponding percent changes were -77.59% (95% CIs: -86.20 to -68.87) and -59.7% (95% CIs: -80.3 to -39.1, p <0.001) **(Figure 1)**.

**Figure 1.**
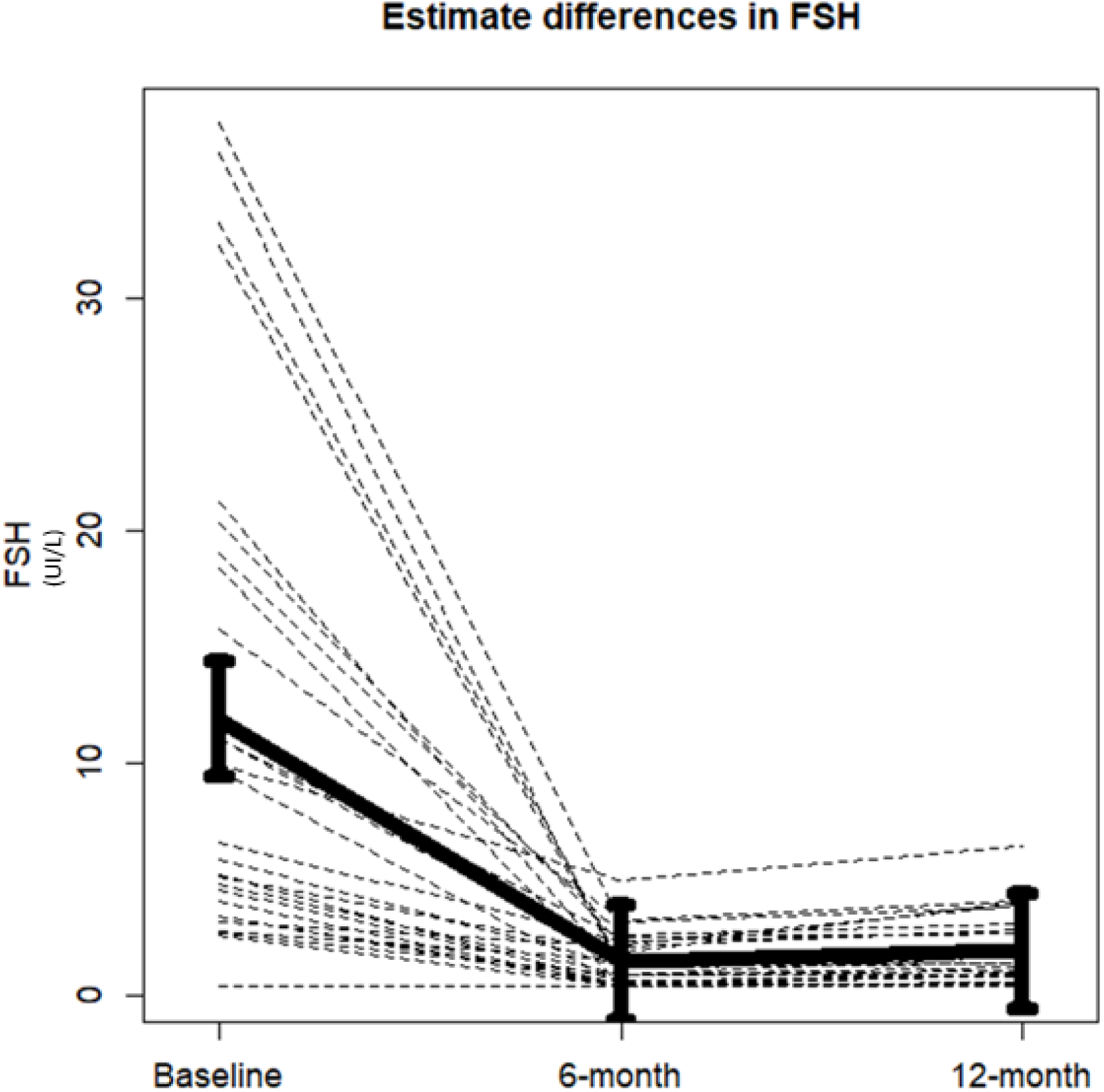
Changes in mean follicle-stimulating hormone (FSH) serum levels from baseline to 6 and 12 months of degarelix treatment.

Data on changes in body composition parameters, BMD and bone turnover markers have been published elsewhere (Palumbo, et al. 2021; Palumbo, et al. 2022; Dalla Volta et al. 2023). Correlations between FSH changes and changes in body composition parameters, BMD and bone turnover markers were not statistically significant, so these data are not presented.

### Relationship between FSH serum levels and body composition, BMD and bone turnover markers at baseline

At baseline conditions no relationship was found between FSH serum levels and the following DXA-derived parameters: BMD at left hip and spine, total FBM, total LBM, lean mass at arms and legs, ALMI, ALMI/FBM ratio and android/gynoid ratio. No correlation was also found between FSH serum levels and serum levels of either ALP or CTX. A direct relationship, although not significant, was found between FSH serum levels and FBM at arms (r=0.36) and legs (r=0.33) (**Table 1**).

**Table 1.** Baseline relationships between FSH serum levels and BMD, bone turnover markers and body composition parameters.

|  | FSH |  |
| --- | --- | --- |
|  | Correlation coefficient | p |
| TOT BMD | .240 | .218 |
| BMD LEFT HIP | .229 | .240 |
| BMD L2-L4 | .148 | .453 |
| CTX | -.115 | .560 |
| BONE ALP | .001 | .997 |
| TOT Fat (g) | .274 | .158 |
| Arm Fat (g) | .363 | .058 |
| Leg Fat (g) | .330 | .087 |
| Head Fat (g) | .228 | .244 |
| Trunk Fat (g) | .249 | .202 |
| TOT Lean (g) | .216 | .270 |
| Arm Lean (g) | .166 | .400 |
| Leg Lean (g) | .197 | .315 |
| Head Lean (g) | .250 | .199 |
| Trunk Lean (g) | .190 | .333 |
| ALMI (Appendicular Lean/Ht<sup>2</sup>) | .269 | .167 |
| ALMI/FBM | -.184 | .349 |
| Android/Gynoid Ratio | -.024 | .902 |

FSH: follicle-stimulating hormone; TOT: total; BMD: bone mineral density; CTX: C-terminal telopeptide of type I collagen; ALP: alkaline phosphatase; ALMI: appendicular lean mass index; Ht<sup>2</sup>: height squared; FBM: fat body mass. Data are Spearman R.

### Relationship between FSH serum levels and body composition, BMD and bone turnover markers after 12 months of degarelix treatment

The correlation between circulating FSH levels at 6 and 12 months, bone turnover markers evaluated at the same time, BMD and body composition parameters assessed after 12 months is shown in **Table 2**. FSH serum levels assessed after 6 months of degarelix administration showed a direct relationship with total FBM (r=0.49, p=0.008) and with fat mass evaluated at arms (r=0.53, p=0.004), trunk (r=0.48, p=0.009) and legs (r=0.45, p=0.015). By contrast, FSH serum levels after 6 months of treatment showed an inverse relationship with ALMI/FBM ratio (r=-0.58, p=0.001). FSH measured after 12 months maintained a significant relationship with total FBM (r=0.52, p=0.006) (**Figure 2A**), fat mass at arms (r=0.54, p=0.004) (**Figure 2B**) and fat mass at trunk (r=0.45, p=0.018) (**Figure 2C**). The relationship between FSH and fat mass at legs lost the statistical significance (r=0.33, p=0.089), while the inverse relationship with ALMI/FBM ratio was confirmed (r=0.64, p=0.001) (**Figure 2D**).

**Figure 2.**
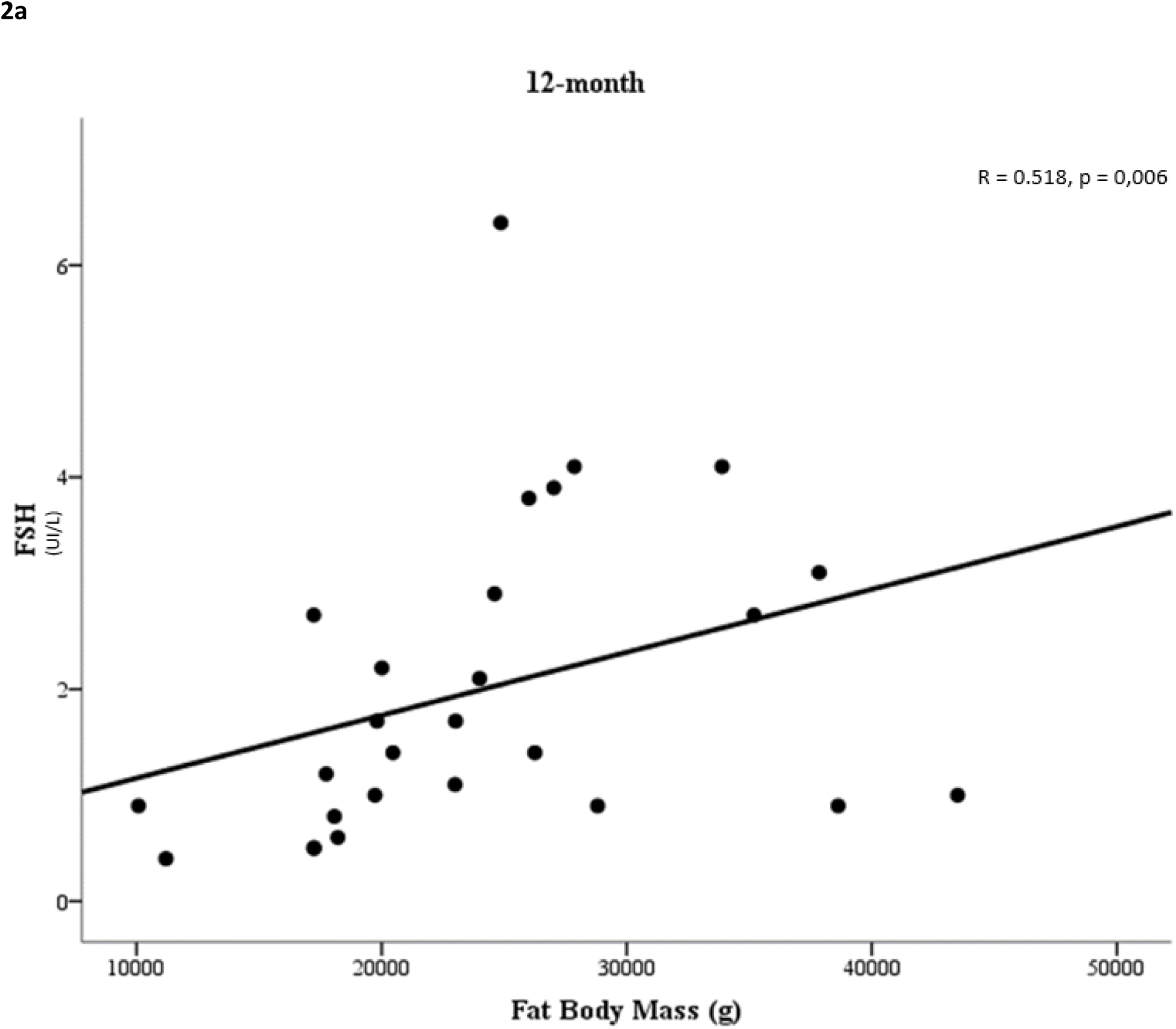
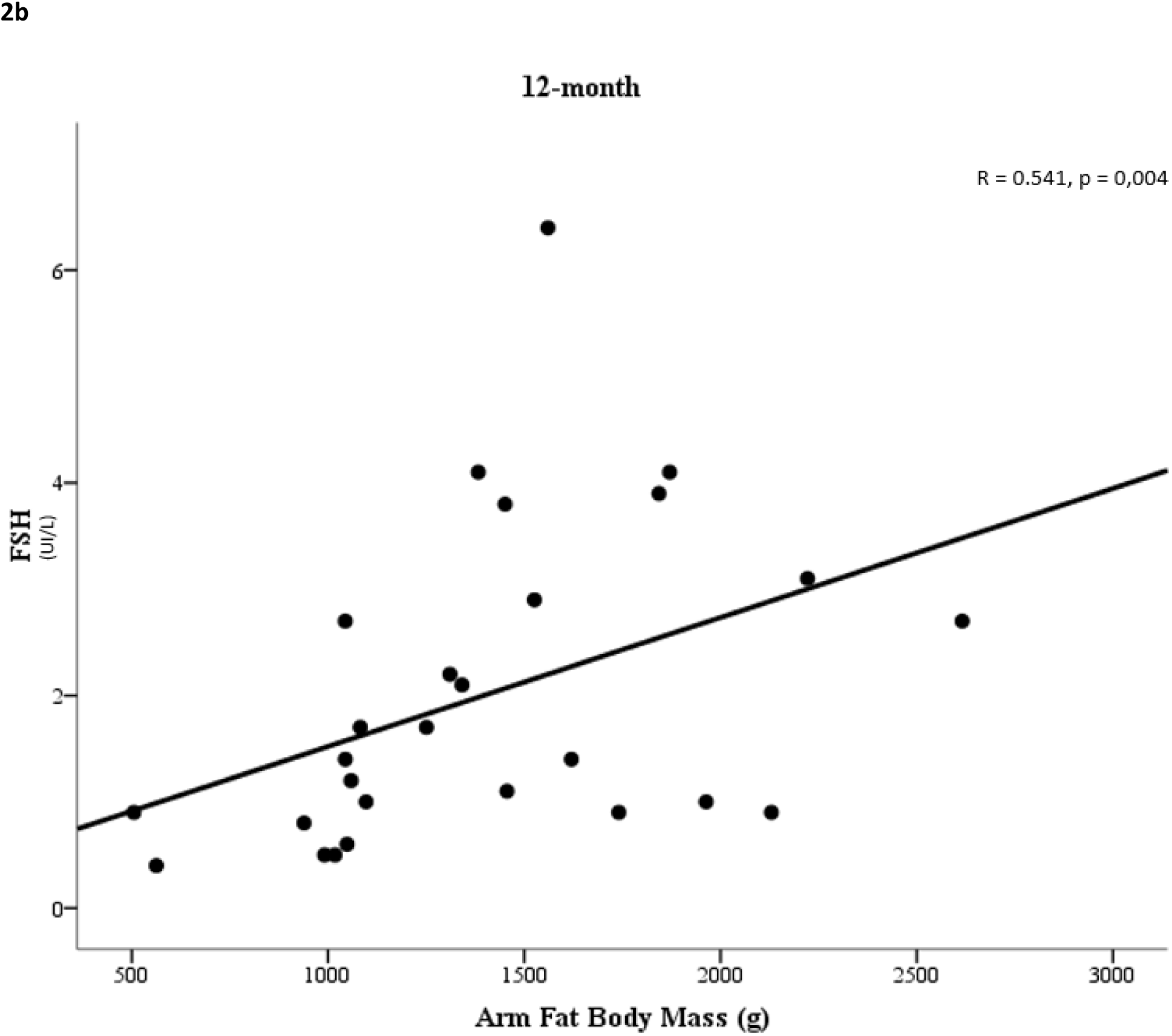
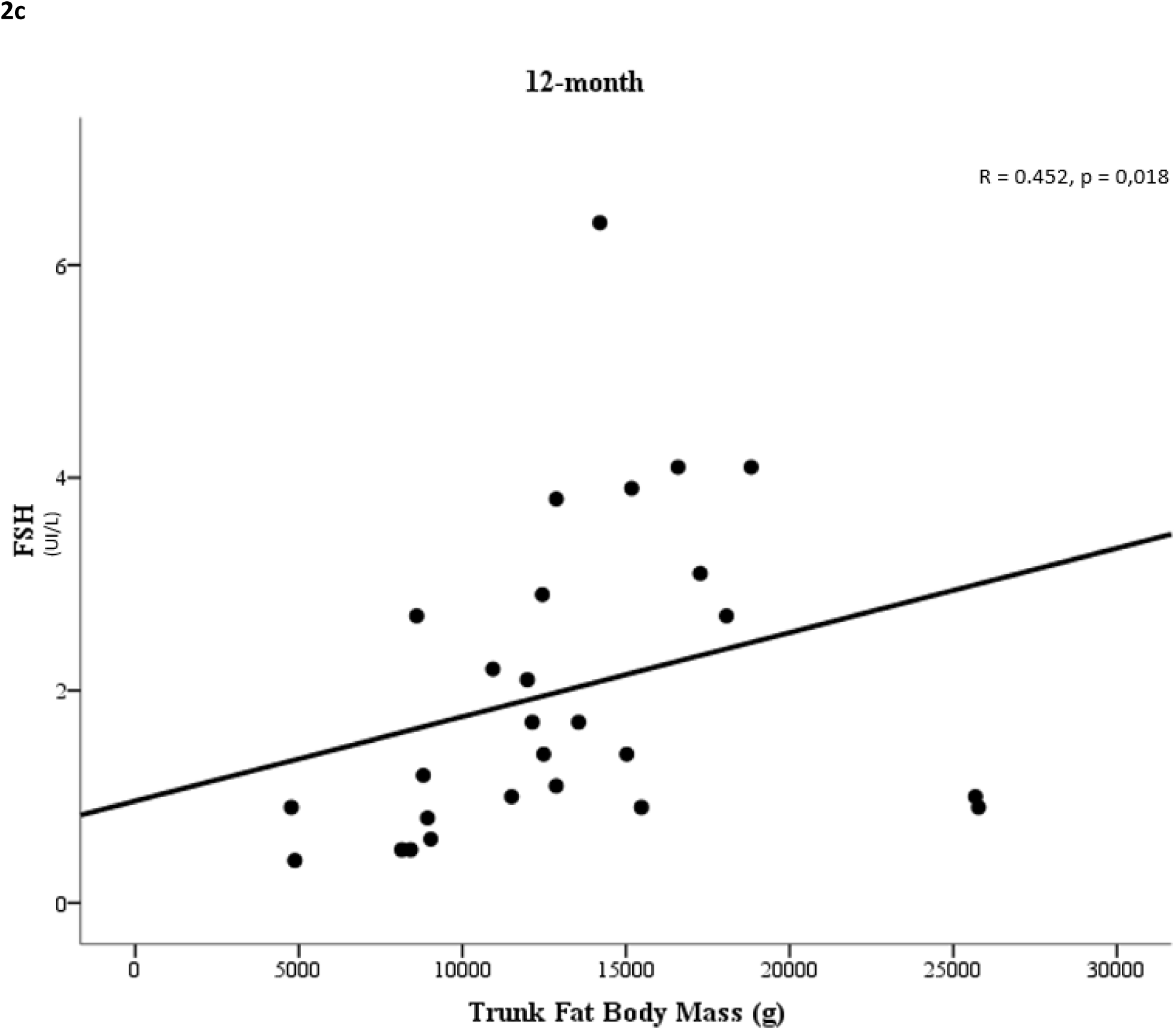
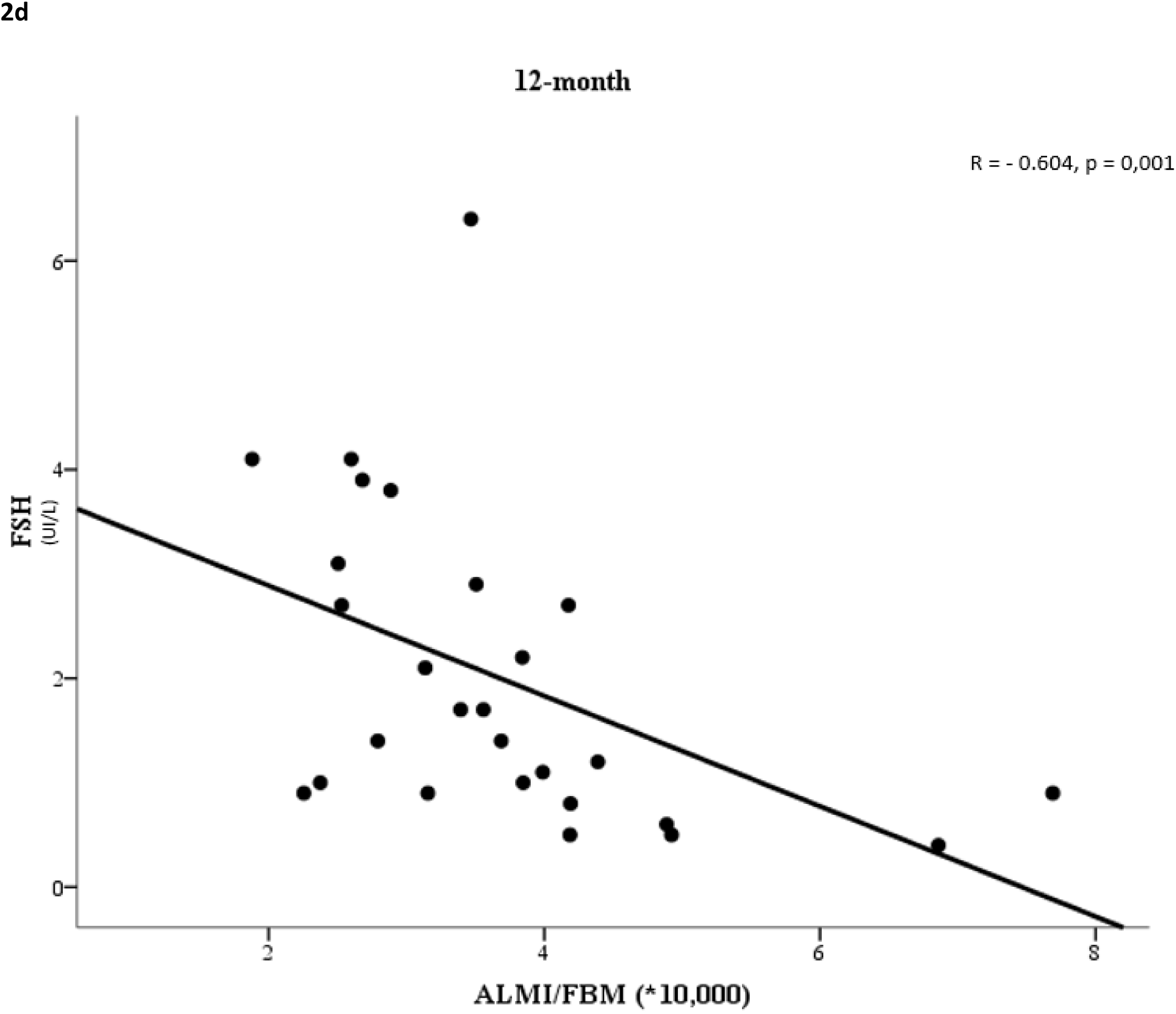
Relationship between follicle-stimulating hormone (FSH) serum levels and fat body mass (FBM) (Fig. 2a), arm fat body mass (Fig. 2b), trunk fat body mass (Fig. 2c) and appendicular lean mass index (ALMI)/FBM ratio (Fig. 2d) after 12-months degarelix treatment.

**Table 2.** Relationships between FSH serum levels and BMD, bone turnover markers and body composition.

|  | FSH at 6 months |  | FSH at 12 months |  |
| --- | --- | --- | --- | --- |
|  | Correlation coefficient | p | Correlation coefficient | p |
| TOT BMD | -.039 | .843 | -.107 | .595 |
| BMD L2-L4 | .102 | .604 | -.040 | .844 |
| BMD LEFT HIP | .030 | .881 | -.044 | .829 |
| CTX | .051 | .796 | .055 | .785 |
| BONE ALP | .174 | .376 | .227 | .256 |
| TOT Fat (g) | **.490** | **.008** | **.518** | **.006** |
| Arm Fat (g) | **.532** | **.004** | **.541** | **.004** |
| Leg Fat (g) | **.454** | **.015** | .328 | .089 |
| Head Fat (g) | .187 | .341 | .093 | .644 |
| Trunk Fat (g) | **.483** | **.009** | **.452** | **.018** |
| TOT Lean (g) | .029 | .885 | -.036 | .859 |
| Arm Lean (g) | .041 | .837 | .028 | .889 |
| Leg Lean (g) | -.176 | .371 | -.285 | .149 |
| Head Lean (g) | .086 | .663 | -.006 | .977 |
| Trunk Lean (g) | .161 | .414 | .021 | .916 |
| ALMI (Appendicular Lean/Ht<sup>2</sup>) | -.229 | .241 | -.265 | .181 |
| ALMI/FBM | **-.581** | **.001\*** | **-.604** | **.001\*** |
| Android/Gynoid Ratio | .142 | .472 | .231 | .246 |

FSH: follicle-stimulating hormone; TOT: total; BMD: bone mineral density; CTX: C-terminal telopeptide of type I collagen; ALP: alkaline phosphatase; ALMI: appendicular lean mass index; Ht<sup>2</sup>: height squared; FBM: fat body mass.
Data are Spearman R.
\*Still significant after Bonferroni correction.

## Discussion

ADT in PC patients leads to an increase in fat mass and to a decrease in lean mass, thus increasing the risk of sarcopenic obesity. In addition, ADT has also a negative impact on skeletal fragility, which increases the risk of fractures. During the recent years, there has been consistent evidence that alterations in body composition could contribute to determine skeletal fragility in patients under ADT, such as well demonstrated in other forms of male hypogonadism (Vena et al. 2022). However, the determinants of altered body composition in male hypogonadism and in ADTs have not been completely defined. As a matter of fact, low testosterone values do not seem to be the only determinant of sarcopenic obesity in males with hypogonadism. In this context, FSH values might play a role. In fact, amongst its extragonadal effects, FSH directly affects adipogenesis, lean mass and bone turnover (Liu, et al. 2017; Rojekar, et al. 2023; Zaidi, et al. 2023; Gera, et al. 2022).

As demonstrated in other studies prospectively evaluating circulating FSH levels after treatment with Luteinizing Hormone-Releasing Hormone (LHRH) agonists and antagonists (Klotz et al. 2008), degarelix administration consistently reduced FSH values. Therefore, it is reasonable to hypothesize that this hormone is involved in the mechanisms that underlie ADT impact on body composition and skeletal fragility.

This ancillary analysis of the BLADE study shows that FSH serum levels correlated with fat mass in basal conditions, albeit not significantly. However, either total fat mass and fat mass in the upper limbs and trunk significantly correlated with FSH values after 12 months of degarelix therapy.

Even more interesting, when assessed after degarelix, FSH levels inversely correlated with ALMI/FBM ratio, a parameter that identifies patients at high risk for sarcopenic obesity. These data suggest that sexual hormones counteract FSH effect on body composition at baseline conditions, whereas this effect becomes more evident after ADT.

It is noteworthy that FSH values and body composition parameters after 12 months of degarelix therapy achieved a significant correlation, despite the narrow range of FSH levels (**Figure 1**). According to these data, small changes in FSH can result in significant changes in body composition in a condition of androgen and estrogen deprivation.

It is known that one of the most noticeable differences of LHRH-antagonists compared to LHRH-agonists are FSH levels during therapy, which are mildly higher with LHRH-agonists compared to LHRH-antagonists. According to our data, PC patients treated with LHRH-antagonists may experience less body composition changes due to lower FSH residual values (Klotz, et al. 2008; Margel et al. 2019; Abufaraj et al. 2021).

Unlike other settings, FSH values before and after degarelix did not correlate with either BMD or bone turnover markers.

To our knowledge, this is the first study showing an association between circulating FSH levels and body composition in PC patients undergoing ADT. However, this study suffers from all the limitations of ancillary studies. Therefore, the results obtained are to be considered as hypothesis-generating and cannot be generalized.

In conclusion, the findings of this study suggest that FSH serum levels after ADT could impact body composition - fat body mass in particular - in PC patients undergoing ADT.

Of course further investigation is required to establish the association between FSH serum levels, measured during ADT, and sarcopenic obesity risk in PC patients. However, this is an area of great interest.

FSH could be a promising marker in monitoring PC patients during ADT in order to identify those who are more vulnerable and at greater risk of cardiovascular complications.

## Data Availability

Data that support the findings of this study are available on request from the corresponding author, Alberto Dalla Volta

## Statements

### Ethical approval

the study was carried out in accordance with the Declaration of Helsinki Principles and Good Clinical Practices and was approved by the Ethics Committee of Brescia (approval numberNP2540). All patients provided a written informed consent.

## Acknowledgements

the authors are grateful to the following associations: AOB (Associazione Oncologica Bresciana) and FIRM-Onlus (Fondazione Internazionale di Ricerca in Medicina Onlus) for the continuous support.

---

**CONSORT DIAGRAM (source data 123547_0_data_set_3007640_s1333z.xls)**

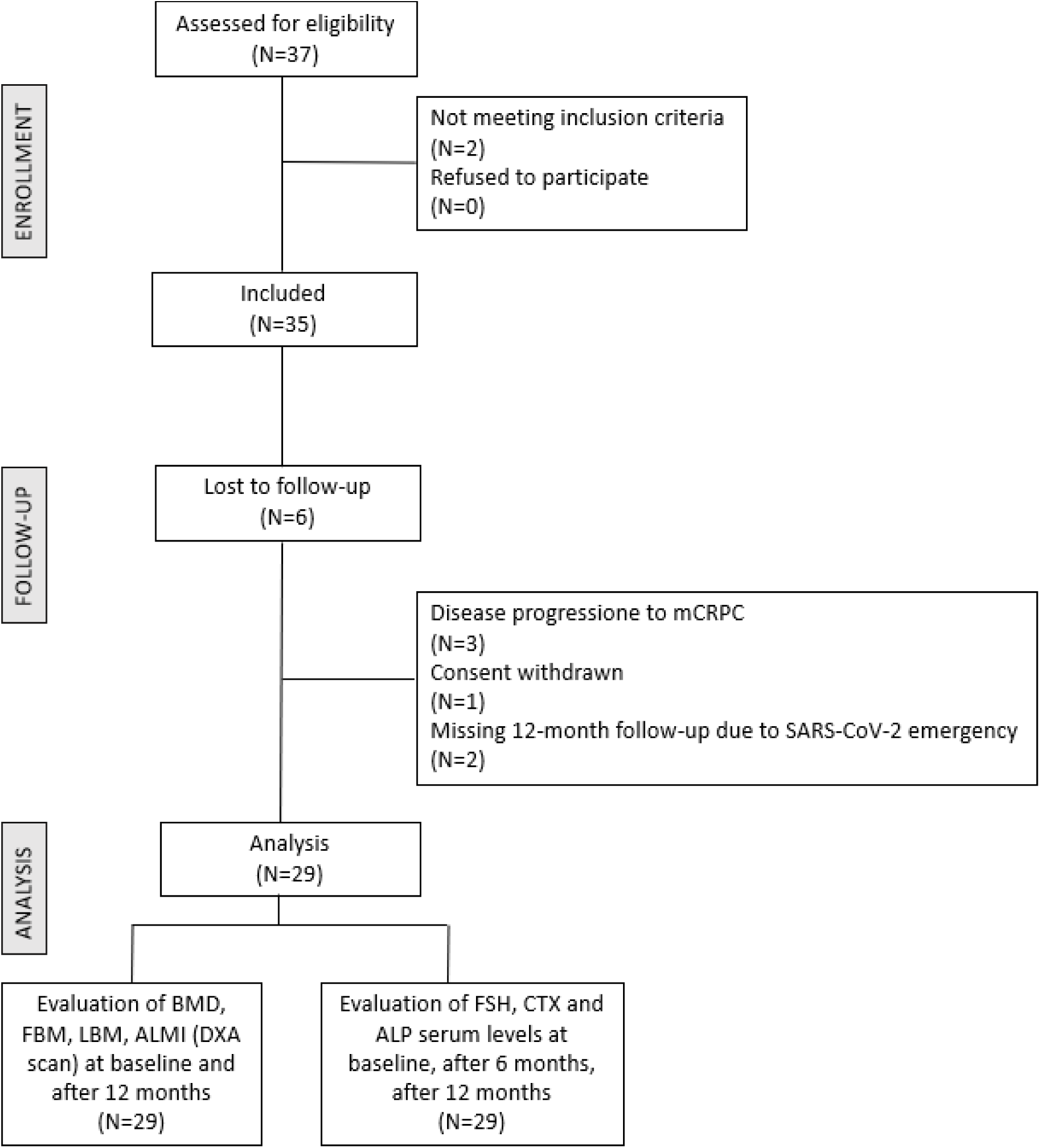

## References

1. Abufaraj, M., T. Iwata, S. Kimura, A. Haddad, H. Al-Ani, L. Abusubaih, M. Moschini, A. Briganti, P. I. Karakiewicz, and S. F. Shariat. 2021. “Differential Impact of Gonadotropin-Releasing Hormone Antagonist Versus Agonist on Clinical Safety and Oncologic Outcomes on Patients with Metastatic Prostate Cancer: A Meta-Analysis of Randomized Controlled Trials.” Eur Urol 79, no. 1 (Jan): 44–53. 10.1016/j.eururo.2020.06.002.

2. Buckinx, F., F. Landi, M. Cesari, R. A. Fielding, M. Visser, K. Engelke, S. Maggi, E. Dennison, N. M. Al-Daghri, S. Allepaerts, J. Bauer, I. Bautmans, M. L. Brandi, O. Bruyère, T. Cederholm, F. Cerreta, A. Cherubini, C. Cooper, A. Cruz-Jentoft, E. McCloskey, B. Dawson-Hughes, J. M. Kaufman, A. Laslop, J. Petermans, J. Y. Reginster, R. Rizzoli, S. Robinson, Y. Rolland, R. Rueda, B. Vellas, and J. A. Kanis. 2018. “Pitfalls in the Measurement of Muscle Mass: A Need for a Reference Standard.” J Cachexia Sarcopenia Muscle 9, no. 2 (04): 269–278. 10.1002/jcsm.12268.

3. Celestino Catão Da Silva, D., A. Nogueira De Almeida Vasconcelos, J. Cleto Maria Cerqueira, D. De Oliveira Cipriano Torres, A. C. Oliveira Dos Santos, H. De Lima Ferreira Fernandes Costa, and L. O. Bregieiro Fernandes Costa. 2013. “Endogenous Sex Hormones Are Not Associated with Subclinical Atherosclerosis in Menopausal Women.” Minerva Ginecol 65, no. 3 (Jun): 297–302.

4. Cornford, P., J. Bellmunt, M. Bolla, E. Briers, M. De Santis, T. Gross, A. M. Henry, S. Joniau, T. B. Lam, M. D. Mason, H. G. van der Poel, T. H. van der Kwast, O. Rouvière, T. Wiegel, and N. Mottet. 2017. “Eau-Estro-Siog Guidelines on Prostate Cancer. Part Ii: Treatment of Relapsing, Metastatic, and Castration-Resistant Prostate Cancer.” Eur Urol 71, no. 4 (04): 630–642. 10.1016/j.eururo.2016.08.002.

5. Dalla Volta, A., C. Palumbo, S. Zamboni, G. Mazziotti, L. Triggiani, M. Zamparini, F. Maffezzoni, L. Rinaudo, M. Bergamini, N. Di Meo, I. Caramella, F. Valcamonico, P. Borghetti, A. Guerini, D. Farina, A. Antonelli, C. Simeone, and A. Berruti. 2023. “Heterogeneity in Regional Changes in Body Composition Induced by Androgen Deprivation Therapy in Prostate Cancer Patients: Potential Impact on Bone Health-the Blade Study.” J Endocrinol Invest (Jul 17). 10.1007/s40618-023-02150-z.

6. El Khoudary, S. R., N. Santoro, H. Y. Chen, P. G. Tepper, M. M. Brooks, R. C. Thurston, I. Janssen, S. D. Harlow, E. Barinas-Mitchell, F. Selzer, C. A. Derby, E. A. Jackson, D. McConnell, and K. A. Matthews. 2016. “Trajectories of Estradiol and Follicle-Stimulating Hormone over the Menopause Transition and Early Markers of Atherosclerosis after Menopause.” Eur J Prev Cardiol 23, no. 7 (May): 694–703. 10.1177/2047487315607044.

7. Gera, S., T. C. Kuo, A. A. Gumerova, F. Korkmaz, D. Sant, V. DeMambro, K. Sudha, A. Padilla, G. Prevot, J. Munitz, A. Teunissen, M. M. T. van Leent, T. G. J.M Post, J. C. Fernandes, J. Netto, F. Sultana, E. Shelly, S. Rojekar, P. Kumar, L. Cullen, J. Chatterjee, A. Pallapati, S. Miyashita, H. Kannangara, M. Bhongade, P. Sengupta, K. Ievleva, V. Muradova, R. Batista, C. Robinson, A. Macdonald, S. Hutchison, M. Saxena, M. Meseck, J. Caminis, J. Iqbal, M. I. New, V. Ryu, S. M. Kim, J. J. Cao, N. Zaidi, Z. A. Fayad, D. Lizneva, C. J. Rosen, T. Yuen, and M. Zaidi. 2022. “Fsh-Blocking Therapeutic for Osteoporosis.” Elife 11 (Sep 20). 10.7554/eLife.78022.

8. Guligowska, A., Z. Chrzastek, M. Pawlikowski, M. Pigłowska, H. Pisarek, K. Winczyk, and T. Kostka. 2021. “Gonadotropins at Advanced Age - Perhaps They Are Not So Bad? Correlations between Gonadotropins and Sarcopenia Indicators in Older Adults.” Front Endocrinol (Lausanne) 12: 797243. 10.3389/fendo.2021.797243.

9. Klotz, L., L. Boccon-Gibod, N. D. Shore, C. Andreou, B. E. Persson, P. Cantor, J. K. Jensen, T. K. Olesen, and F. H. Schröder. 2008. “The Efficacy and Safety of Degarelix: A 12-Month, Comparative, Randomized, Open-Label, Parallel-Group Phase Iii Study in Patients with Prostate Cancer.” BJU Int 102, no. 11 (Dec): 1531–8. 10.1111/j.1464-410X.2008.08183.x.

10. Liu, P., Y. Ji, T. Yuen, E. Rendina-Ruedy, V. E. DeMambro, S. Dhawan, W. Abu-Amer, S. Izadmehr, B. Zhou, A. C. Shin, R. Latif, P. Thangeswaran, A. Gupta, J. Li, V. Shnayder, S. T. Robinson, Y. E. Yu, X. Zhang, F. Yang, P. Lu, Y. Zhou, L. L. Zhu, D. J. Oberlin, T. F. Davies, M. R. Reagan, A. Brown, T. R. Kumar, S. Epstein, J. Iqbal, N. G. Avadhani, M. I. New, H. Molina, J. B. van Klinken, E. X. Guo, C. Buettner, S. Haider, Z. Bian, L. Sun, C. J. Rosen, and M. Zaidi. 2017. “Blocking Fsh Induces Thermogenic Adipose Tissue and Reduces Body Fat.” Nature 546, no. 7656 (06 01): 107–112. 10.1038/nature22342.

11. Lizneva, D., A. Rahimova, S. M. Kim, I. Atabiekov, S. Javaid, B. Alamoush, C. Taneja, A. Khan, L. Sun, R. Azziz, T. Yuen, and M. Zaidi. 2019. “Fsh Beyond Fertility.” Front Endocrinol (Lausanne) 10: 136. 10.3389/fendo.2019.00136.

12. Margel, D., A. Peer, Y. Ber, L. Shavit-Grievink, T. Tabachnik, S. Sela, G. Witberg, J. Baniel, D. Kedar, W. C. M. Duivenvoorden, E. Rosenbaum, and J. H. Pinthus. 2019. “Cardiovascular Morbidity in a Randomized Trial Comparing Gnrh Agonist and Gnrh Antagonist among Patients with Advanced Prostate Cancer and Preexisting Cardiovascular Disease.” J Urol 202, no. 6 (Dec): 1199–1208. 10.1097/JU.0000000000000384.

13. Mattick, L. J., J. W. Bea, L. Singh, K. M. Hovey, H. R. Banack, J. Wactawski-Wende, J. E. Manson, J. L. Funk, and H. M. Ochs-Balcom. 2022. “Serum Follicle-Stimulating Hormone and 5-Year Change in Adiposity in Healthy Postmenopausal Women.” J Clin Endocrinol Metab 107, no. 8 (07 14): e3455–e3462. 10.1210/clinem/dgac238.

14. Mottet, N., J. Bellmunt, M. Bolla, E. Briers, M. G. Cumberbatch, M. De Santis, N. Fossati, T. Gross, A. M. Henry, S. Joniau, T. B. Lam, M. D. Mason, V. B. Matveev, P. C. Moldovan, R. C. N. van den Bergh, T. Van den Broeck, H. G. van der Poel, T. H. van der Kwast, O. Rouvière, I. G. Schoots, T. Wiegel, and P. Cornford. 2017. “Eau-Estro-Siog Guidelines on Prostate Cancer. Part 1: Screening, Diagnosis, and Local Treatment with Curative Intent.” Eur Urol 71, no. 4 (04): 618–629. 10.1016/j.eururo.2016.08.003.

15. Munir, J. A., H. Wu, K. Bauer, J. Bindeman, C. Byrd, I. M. Feuerstein, T. C. Villines, and A. J. Taylor. 2012. “The Perimenopausal Atherosclerosis Transition: Relationships between Calcified and Noncalcified Coronary, Aortic, and Carotid Atherosclerosis and Risk Factors and Hormone Levels.” Menopause 19, no. 1 (Jan): 10–5. 10.1097/gme.0b013e318221bc8d.

16. Palumbo, C., A. Antonelli, L. Triggiani, A. Dalla Volta, F. Maffezzoni, S. Zamboni, P. Borghetti, L. Rinaudo, F. Valcamonico, R. Maroldi, S. M. Magrini, C. Simeone, A. Berruti, and Collaborators. 2021. “Changes in Body Composition and Lipid Profile in Prostate Cancer Patients without Bone Metastases Given Degarelix Treatment: The Blade Prospective Cohort Study.” Prostate Cancer Prostatic Dis 24, no. 3 (09): 852–859. 10.1038/s41391-021-00345-0.

17. Palumbo, C., A. Dalla Volta, S. Zamboni, G. Mazziotti, M. Zamparini, L. Triggiani, P. Borghetti, F. Maffezzoni, R. Bresciani, L. Rinaudo, F. Valcamonico, D. Farina, S. M. Magrini, A. Antonelli, C. Simeone, and A. Berruti. 2022. “Effect of Degarelix Administration on Bone Health in Prostate Cancer Patients without Bone Metastases. The Blade Study.” J Clin Endocrinol Metab (Aug 16). 10.1210/clinem/dgac489.

18. Perneger, T. V. 1998. “What’s Wrong with Bonferroni Adjustments.” BMJ 316, no. 7139 (Apr 18): 1236–8. 10.1136/bmj.316.7139.1236.

19. Rojekar, S., A. R. Pallapati, J. Gimenez-Roig, F. Korkmaz, F. Sultana, D. Sant, C. M. Haeck, A. Macdonald, S. M. Kim, C. J. Rosen, O. Barak, M. Meseck, J. Caminis, D. Lizneva, T. Yuen, and M. Zaidi. 2023. “Development and Biophysical Characterization of a Humanized Fsh-Blocking Monoclonal Antibody Therapeutic Formulated at an Ultra-High Concentration.” Elife 12 (Jun 19). 10.7554/eLife.88898.

20. Senapati, S., C. R. Gracia, E. W. Freeman, M. D. Sammel, H. Lin, C. Kim, R. J. Schwab, and G. W. Pien. 2014. “Hormone Variations Associated with Quantitative Fat Measures in the Menopausal Transition.” Climacteric 17, no. 2 (Apr): 183–90. 10.3109/13697137.2013.845876.

21. Sowers, M., H. Zheng, K. Tomey, C. Karvonen-Gutierrez, M. Jannausch, X. Li, M. Yosef, and J. Symons. 2007. “Changes in Body Composition in Women over Six Years at Midlife: Ovarian and Chronological Aging.” J Clin Endocrinol Metab 92, no. 3 (Mar): 895–901. 10.1210/jc.2006-1393.

22. Sun, L., Y. Peng, A. C. Sharrow, J. Iqbal, Z. Zhang, D. J. Papachristou, S. Zaidi, L. L. Zhu, B. B. Yaroslavskiy, H. Zhou, A. Zallone, M. R. Sairam, T. R. Kumar, W. Bo, J. Braun, L. Cardoso-Landa, M. B. Schaffler, B. S. Moonga, H. C. Blair, and M. Zaidi. 2006. “Fsh Directly Regulates Bone Mass.” Cell 125, no. 2 (Apr 21): 247–60. 10.1016/j.cell.2006.01.051.

23. Thurston, R. C., M. R. Sowers, B. Sternfeld, E. B. Gold, J. Bromberger, Y. Chang, H. Joffe, C. J. Crandall, L. E. Waetjen, and K. A. Matthews. 2009. “Gains in Body Fat and Vasomotor Symptom Reporting over the Menopausal Transition: The Study of Women’s Health across the Nation.” Am J Epidemiol 170, no. 6 (Sep 15): 766–74. 10.1093/aje/kwp203.

24. Vena, W., F. Carrone, A. Delbarba, O. Akpojiyovbi, L. C. Pezzaioli, P. Facondo, C. Cappelli, L. Leonardi, L. Balzarini, D. Farina, A. Pizzocaro, A. G. Lania, G. Mazziotti, and A. Ferlin. 2022. “Body Composition, Trabecular Bone Score and Vertebral Fractures in Subjects with Klinefelter Syndrome.” J Endocrinol Invest (Aug 28). 10.1007/s40618-022-01901-8.

25. Visser, M., T. Fuerst, T. Lang, L. Salamone, and T. B. Harris. 1999. “Validity of Fan-Beam Dual-Energy X-Ray Absorptiometry for Measuring Fat-Free Mass and Leg Muscle Mass. Health, Aging, and Body Composition Study--Dual-Energy X-Ray Absorptiometry and Body Composition Working Group.” J Appl Physiol (1985) 87, no. 4 (Oct): 1513–20. 10.1152/jappl.1999.87.4.1513.

26. Zaidi, M., S. M. Kim, M. Mathew, F. Korkmaz, F. Sultana, S. Miyashita, A. A. Gumerova, T. Frolinger, O. Moldavski, O. Barak, A. Pallapati, S. Rojekar, J. Caminis, Y. Ginzburg, V. Ryu, T. F. Davies, D. Lizneva, C. J. Rosen, and T. Yuen. 2023. “Bone Circuitry and Interorgan Skeletal Crosstalk.” Elife 12 (Jan 19). 10.7554/eLife.83142.

